# Differential benefit of adjuvant everolimus according to endocrine therapy backbone in the randomized UNIRAD trial

**DOI:** 10.1101/2024.10.01.24314713

**Authors:** Mathilde Saint-Ghislain, Sylvie Chabaud, Florence Dalenc, Djelila Allouache, David Cameron, Mathilde Martinez, Julien Grenier, Philippe Barthelemy, Murray Brunt, Laure Kaluzinski, Audrey Mailliez, Eric Legouffe, Anne-Claire Hardy-Bessard, Sylvie Giacchetti, Marie-Ange Mouret-Reynier, Jean-Luc Canon, Judith Bliss, Jérôme Lemonnier, Fabrice Andre, Thomas Bachelot, Paul Cottu

## Abstract

**Background:** The randomized, double-blind UNIRAD trial evaluating the addition of 2 years of everolimus to endocrine therapy in patients with high-risk, early luminal breast cancer failed to demonstrate a benefit. We report the subgroup analyses.

**Patients and Methods:** We randomized 1278 patients in a 1:1 ratio to receive 2 years of placebo or everolimus, added to endocrine therapy for up to 4 years after initiation. Randomization was stratified by endocrine therapy agent, prior adjuvant versus neoadjuvant therapy, progesterone receptor expression, and lymph node involvement. Subgroup analyses by each stratification factor were prespecified. Post hoc analyses were performed according to menopausal status and age. We also analyzed treatment adherence.

**Results:** We observed a limited trend toward more favorable prognostic features in tamoxifen-treated patients, with more frequent ER+/PR+ tumors (88.5% vs. 84.1%, p=0.026) and less frequent pN2+ status (39.8% vs. 46%, p=0.032). In premenopausal women, we observed a numerical benefit of everolimus: 3y-DFS was 86% in the placebo group and 90% in the everolimus group [HR=0.76 (95%CI: 0.43-1.34)]. In premenopausal patients treated with tamoxifen (n=153; 12.3%), we observed an even stronger trend in favor of everolimus as 3-year DFS was 84% in the placebo group and 91% in the everolimus group [HR=0.54 (95%CI: 0.28-1.02)]. Early discontinuation of either everolimus or placebo was less frequent in the tamoxifen group than in the AI group: 48.0% vs. 56.9% (p=0.028).

**Conclusions:** The present post-hoc analyses generate hypotheses regarding the interaction between menopausal status, tamoxifen and everolimus in patients with high-risk, ER-positive, HER2-negative early breast cancer. They suggest that tamoxifen alone is no longer the standard of care in high-risk premenopausal patients.

## Introduction

Hormone receptor (HR)-positive, human epidermal growth factor receptor type 2 (HER2)-negative breast cancer accounts for approximately 70% of early invasive breast cancer (EBC) ^1^. Endocrine therapy (ET) is the cornerstone of medical adjuvant therapy and is associated with long-term benefit in both invasive disease-free and overall survival ^2^. In menopausal patients, aromatase inhibitors for at least 5 years are the standard of care^3^. In premenopausal patients, at least five years of tamoxifen has been the standard adjuvant endocrine therapy for decades^4^. More recently, it has been suggested that the combination of any endocrine therapy with ovarian function suppression for 5 years significantly improves invasive disease-free survival over tamoxifen alone, especially in very young patients (<35 years old) and in patients with high-risk features who usually also require adjuvant chemotherapy ^5,6^. Unfortunately, approximately 20% of all patients with ER+, HER2-EBC will relapse in the first 10 years with potentially incurable disease. The long-term risk of recurrence is associated with well-documented prognostic features such as tumor size, lymph node involvement, vascular emboli and tumor proliferation ^7^.

Improving long-term outcomes with adjuvant therapies in these patients, whether premenopausal or not, remains an important unmet medical need. Recently, controversial results have challenged the role of adjuvant CDK4/6 inhibitors, as a means to limit long-term endocrine resistance by targeting the cell cycle machinery ^8^. Based on the results of the Monarch-E trial^8^, abemaciclib was approved by the FDA and the EMA for use in patients with very high-risk early ER+, HER2-BC ^9^. Two adjuvant trials failed to demonstrate any benefit of palbociclib^10^, while results from the NATALEE study evaluating ribociclib in patients with intermediate and high-risk EBC showed a statistically significant improvement of invasive disease-free survival^11^.

Similarly, activation of the phosphatidylinositol-3-kinase (PI3K)/Akt/mammalian target of rapamycin (mTOR) signaling pathway has been described as a mechanism of acquired resistance to endocrine therapy ^12^. In patients with metastatic disease the combination of everolimus and exemestane improved the median progression-free survival (PFS) from 4.1 months to 10.6 months in the BOLERO2 trial^13^. In the TAMRAD trial, the combination of tamoxifen plus everolimus improved the 6-month clinical benefit rate (CBR) from 42% in the tamoxifen arm to 61% in the combination arm ^14^. Within this context and prior to the CDK4/6 inhibitor era, we initiated UNIRAD, a double-blind, multicenter, international randomized trial comparing the combination of adjuvant everolimus plus standard adjuvant endocrine therapy versus placebo plus endocrine therapy in patients with high-risk hormone receptor-positive HER2-negative early breast cancer. The primary results have been reported, suggesting no overall benefit from the addition of 2 years of everolimus to endocrine therapy^15^. The pre-specified subgroup analysis of the UNIRAD trial focused on the endocrine therapy backbone as it appeared to be the only stratification factor with a strong trend toward an association with disease free survival, suggesting that patients treated with tamoxifen may derive a benefit from the addition of everolimus [HR=0.62; 95%CI: 0.369-1.06), p=0.044]^15^. To further elucidate this intriguing finding, we examine in this report the interaction of endocrine therapy and allocation to everolimus with age and menopausal status.

## Patients and Methods

### Patients

Detailed methods have been previously reported^15^. Briefly, patients were enrolled if they were women aged ≥18 years with estrogen receptor-positive, human epidermal growth factor receptor type 2 (HER2)-negative early breast cancer at high risk of recurrence, as defined by ≥4 positive lymph nodes; ≥1 positive lymph node if surgery was performed after neoadjuvant chemotherapy or endocrine therapy administered for ≥3 months; or 1-3 positive lymph nodes at primary surgery and an EPClin® score ≥3.3.

Key non-inclusion criteria were: prior cancer ≤5 years prior to randomization, significantly impaired lung function, known hypersensitivity to mTOR inhibitors, and any uncontrolled medical condition.

The trial was conducted in accordance with good clinical practice, the Declaration of Helsinki, and all local regulations. Written informed consent was obtained from all patients. The trial was registered on clinicaltrials.gov (NCT01805271). It was sponsored and conducted by UNICANCER Research and Development.

### Study design and conduct of treatment

Patients were randomized 1:1 to receive 2 years of placebo or 2 years of everolimus, which was added to ongoing endocrine therapy for up to 4 years after initiation. Randomization was stratified by endocrine therapy agent (tamoxifen +/- luteinizing hormone-releasing hormone [LHRH] agonists vs. AI), prior adjuvant versus neoadjuvant chemotherapy or endocrine therapy, progesterone receptor expression, prior duration of endocrine therapy (≤3 years vs. >3 years), and lymph node involvement (≥4 positive lymph nodes and ≥1 positive lymph node after neoadjuvant treatment vs. 1-3 positive nodes and high EPclin® score). Everolimus dose adjustments have been previously reported ^15^.

### Statistical analysis

The primary endpoint of the core analysis was disease-free survival (DFS) from randomization. The primary analysis was conducted according to the ITT principle. Subgroup analyses according to each stratification factor were prespecified in the statistical analysis plan. For the purpose of this report, post hoc analyses were performed according to menopausal status and age. Data are reported according to the CONSORT 2010 statement^16^.

## Results

### Overall population results

Between June 2013 and March 2020, 1278 patients were randomized, including 641 patients in the placebo arm, and 637 patients in the everolimus arm. The overall population has been previously reported^15^. Briefly, the median age was 54 years (interquartile range, IQR=48-63) and 404 and 838 women were premenopausal (31.6%) and menopausal (65.8%), respectively (unknown 2.5%). The median duration of endocrine therapy treatment at randomization was 15 months (IQR 4.9-29.9). Endocrine therapies included tamoxifen (43.6%), letrozole (31.7%), anastrozole (18.9%), and exemestane (5.4%). Only seven patients (0.5%) received a luteinizing hormone-releasing hormone agonist. In the pre-specified subgroup analysis, endocrine therapy backbone emerged as the only stratification factor with a strong trend toward an association with disease-free survival (p-value for interaction=0.044), suggesting a benefit only in patients receiving tamoxifen^15^.

Other stratification factors such as time to chemotherapy, progesterone receptor expression, duration of endocrine therapy at everolimus initiation or nodal involvement were not significantly associated with DFS. These results have been updated, with a median follow-up of 60.3 months (IQR 42.2 – 71.8), compared to 35.7 months in the original report. In the subgroup of patients receiving tamoxifen, DFS at 60 months was 87% (95%CI, 81 to 91) in the everolimus arm and 80% (95%CI, 74 to 84) in the placebo arm (HR = 0.53; 95%CI 0.33 to 0.85; p=0.0067)(Figure 1A). Conversely, in the AI subgroup, 60-month DFS was 81% (95%CI, 76 to 85) in the everolimus arm and 82% (95%CI, 77 to 86) in the placebo arm (HR = 1.13; 95%CI 0.81 to 1.58; p=0.4736)(Figure 1B). Similar results were observed for distant metastasis-free survival (Figure 1C & 1D).

**Figure 1.**
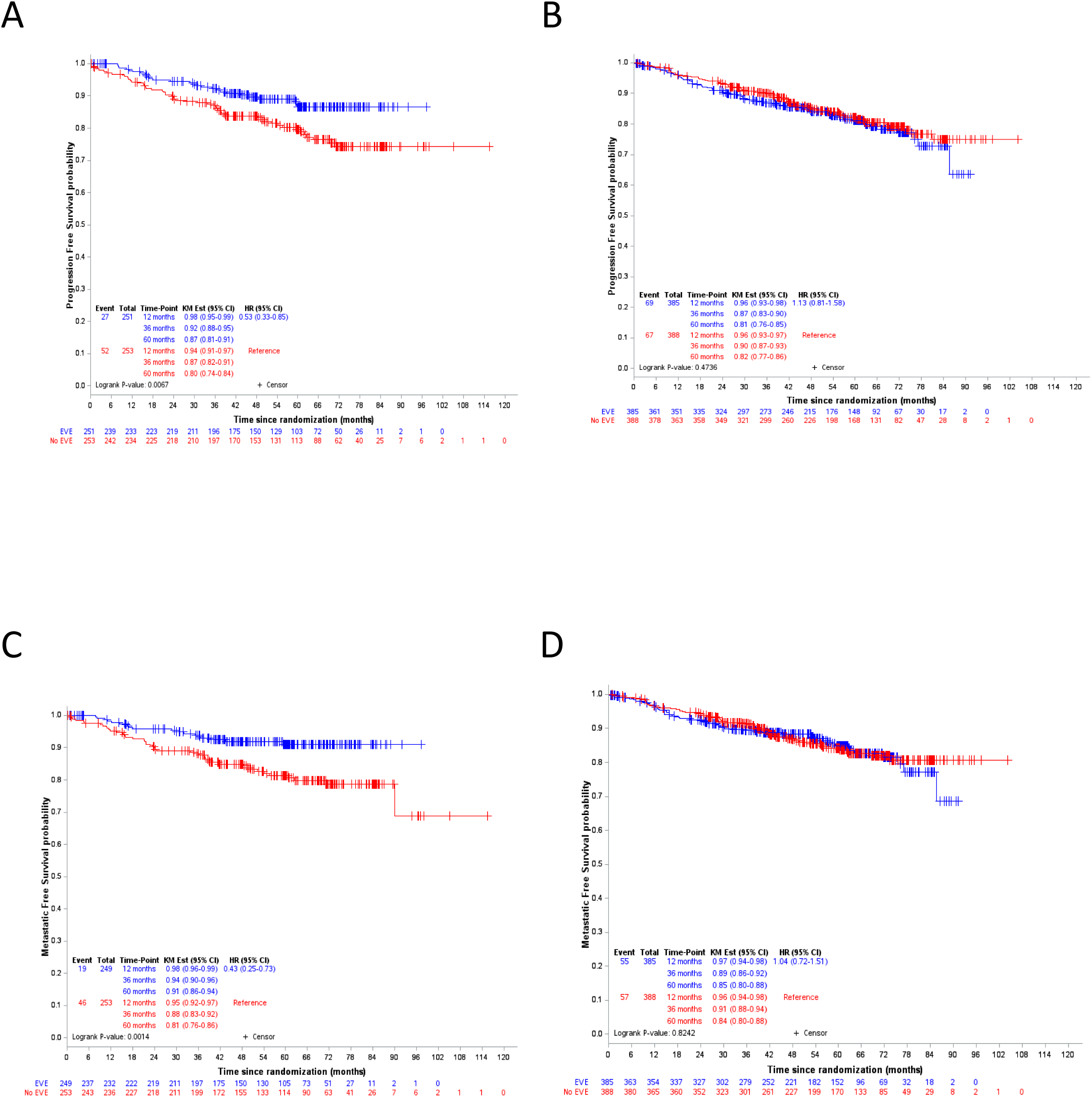
Subgroup analysis of disease-free survival and distant metastasis-free survival according to endocrine therapy backbone. Blue curves indicate the everolimus arm; red curves indicate the placebo arm. (A) DFS in tamoxifen subgroup. (B) DFS in the aromatase inhibitor subgroup. (C) DMFS in the tamoxifen subgroup. (D) DMFS in aromatase inhibitor subgroup. DFS, disease-free survival; DMFS, distant metastasis-free survival; HR, hazard ratio

To further investigate this puzzling interaction, we developed three approaches. First, we looked for potentially different characteristics in these two subsets of patients (i.e., tamoxifen or AI endocrine therapy). Second, we examined the interaction of menopausal status and age with DFS and treatments. Finally, we analyzed the relationship between dosing and compliance with everolimus therapy with either patient characteristics or endocrine therapy treatment.

### Subpopulations

We reanalyzed the study population according to endocrine therapy backbone (Table 1). Overall, no major differences in high-risk characteristics were observed between the 2 groups. However, we observed a limited trend toward more favorable prognostic features in tamoxifen-treated patients, who were more likely to have ER+/PR+ tumors (88.5% vs. 84.1%, exploratory p-value=0.026) and less likely to have pN2+ lymph node involvement (49.6% vs. 57.3%, exploratory p-value=0.022) than aromatase inhibitor-treated patients. As expected, age and menopausal status differed, as patients receiving an aromatase inhibitor were significantly older and more likely to be menopausal.

**Table 1.** Population characteristics according to endocrine therapy.

|  | **Tamoxifen**<br>N=505 | **Aromatase inhibitor**<br>N=773 | **P value** |
| --- | --- | --- | --- |
| **Age**<br>**Median (min-max)** | 47 (28-73) | 60 (31-89) | < 0.001 |
| **Menopause** |  |  | < 0.0001 |
| Yes | 154 (31.7%) | 688 (90.8%) |  |
| No | 332 (68.3%) | 70 (9.2%) |  |
| **missing** | 19 | 15 |  |
| **Tumor characteristics** |  |  |  |
| ER+PR+ | 447(88.5%) | 650 (84.1%) | 0.026 |
| ER+/PR- | 58 (11.5%) | 123 (15.9%) |  |
| ≥4N+ | 250 (49.7%) | 443 (57.5%) | 0.019 |
| 1-3 N+ and EPClin® score ≥ 3.3 | 183 (36.4%) | 229 (29.9%) |  |
| 1-3 N+ after neo-adjuvant treatment | 70 (13.9%) | 98 (12.7%) |  |
| **missing** | 2 | 3 |  |
| **Chemotherapy** |  |  | 0.834 |
| Neoadjuvant | 132 (26.1%) | 198 (25.6%) |  |
| adjuvant | 373 (73.9%) | 575 (74.4%) |  |
| **Duration of endocrine therapy (year)** |  |  | 0.132 |
| 0-1 | 227 (45.0%) | 304 (39.4%) |  |
| 2-3 | 204 (40.5%) | 347 (44.9%) |  |
| >3 | 73 (14.5%) | 121 (15.7%) |  |
| **missing** | 1 | 1 |  |
| **Treatment arm** |  |  | 0.973 |
| Placebo | 253 (50.1%) | 388 (50.2%) |  |
| everolimus | 252 (49.9%) | 385 (49.8%) |  |

### Disease free survival according to endocrine treatment, menopausal status and age

In this post-hoc unplanned subgroup analysis, we examined the interaction between endocrine therapy backbone, menopausal status, and age. In premenopausal women, we observed a non-statistically significant numerical benefit of everolimus: 3y-DFS was 86% (95%CI: 79-91) in the placebo group and 90% (95%CI: 83-94) in the everolimus group [HR=0.76 (95%CI: 0.43-1.34), p=0.3432, Figure 2A], while no difference was observed in menopausal patients: 3y-DFS was 90% (95%CI: 86-93) in the placebo group and 88% (95%CI: 84-91) in the everolimus group [HR=1.04 (95%CI: 0.70-1.55), p=0.8451, Figure 2B]. Interestingly, in premenopausal patients treated with tamoxifen (n=332; 26.7%), we observed an even greater trend in favor of everolimus, as 3-year DFS was 84% (95%CI: 76-89) for the placebo group and 91% (95%CI: 84-95) for the everolimus group [HR=0.54 (95%CI: 0.28-1.02), p=0.0521, Figure 2C]. No benefit of everolimus was observed in menopausal patients treated with tamoxifen (Figure 2D). Conversely, we observed an intriguing potential detrimental effect of everolimus in the subgroup of 72 premenopausal patients treated with an AI, as the 3-year DFS was 95% (95%CI: 68-99) for the control group and 85% (95%CI: 65-94) for the everolimus group [HR=4.78 (95%CI: 0.96-23.82), p=0.0355 Figure 2E], while again no difference was observed in menopausal women treated with an AI (Figure 2F). This small subset of patients had a median age of 47.5 years (range: 31-59; mean: 46 years) and 20 of them had also received tamoxifen as part of their endocrine therapy. Finally, age groups were not associated with any trend, whether considering patients under 45 [HR=0.90, (95%CI 0.45-1.80), p=0.7662] or under ≥45 [HR=0.97, (95%CI 0.67-1.40), p=0.8810] (Supplementary Figure).

**Figure 2.**
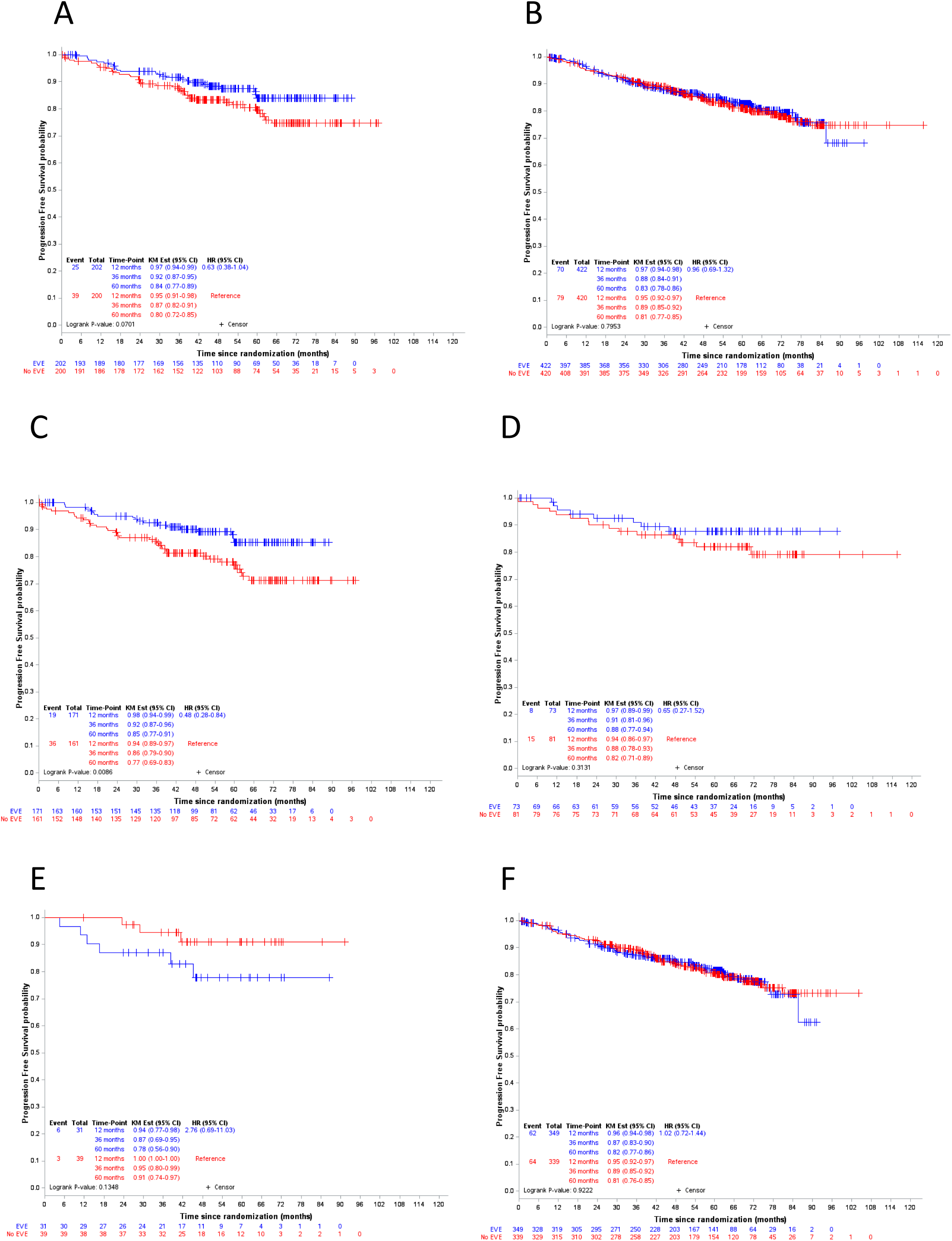
Subgroup analysis of disease-free survival according to menopausal status and endocrine therapy. Blue curves indicate the everolimus arm; red curves indicate the placebo arm. (A) DFS in all non-menopausal patients. (B) DFS in all menopausal patients. (C) DFS in premenopausal patients treated with tamoxifen. (D) DFS in menopausal patients treated with tamoxifen. (E) DFS in premenopausal patients treated with an aromatase inhibitor. (F) DFS in menopausal patients treated with an aromatase inhibitor. DFS, disease-free survival; HR, hazard ratio

### Dose of treatment and adverse events

Finally, to explain these differences, we examined whether dosing and compliance with everolimus therapy might be associated with either patient characteristics or endocrine therapy (Table 2). In the everolimus and tamoxifen group, 84 patients (33.3%) started everolimus at 10 mg and 165 (65.5%) at 5 mg, compared with 35.3% and 61.3%, respectively, in the AI group. In the tamoxifen plus everolimus arm, 245 patients (97.6%) experienced at least one adverse event, including 78 patients (31.1%) with grade 3-5 adverse events. Grade 3-5 adverse events were observed in 41 patients (24.8%) when the everolimus dose was started at 5 mg and in 36 patients (42.9%) when the dose was started at 10 mg. In the AI + everolimus arm, 368 patients (98.4%) experienced at least one AE and 109 (29.1%) experienced a grade 3-5 AE. Similarly, this was observed in 61 patients (25.8%) when everolimus was initiated at 5 mg and in 48 patients (35.3%) at 10 mg. The rate of dose reduction was similar in tamoxifen and AI patients, occurring in 34.9% of the tamoxifen everolimus group and 33.8% of the AI everolimus group. Overall, these results do not suggest a relevant difference in global toxicity profile according to the endocrine therapy partner.

**Table 2.**
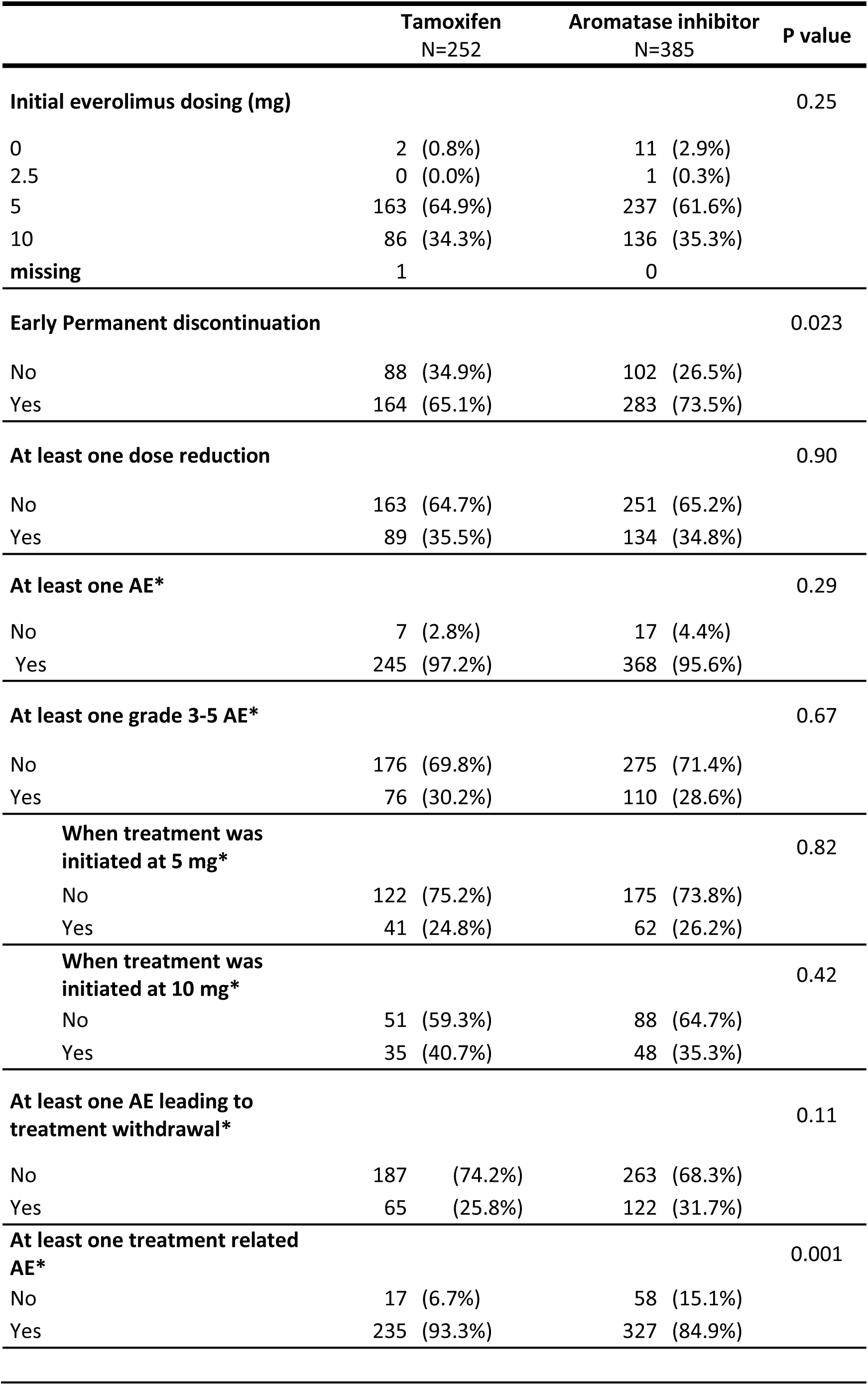
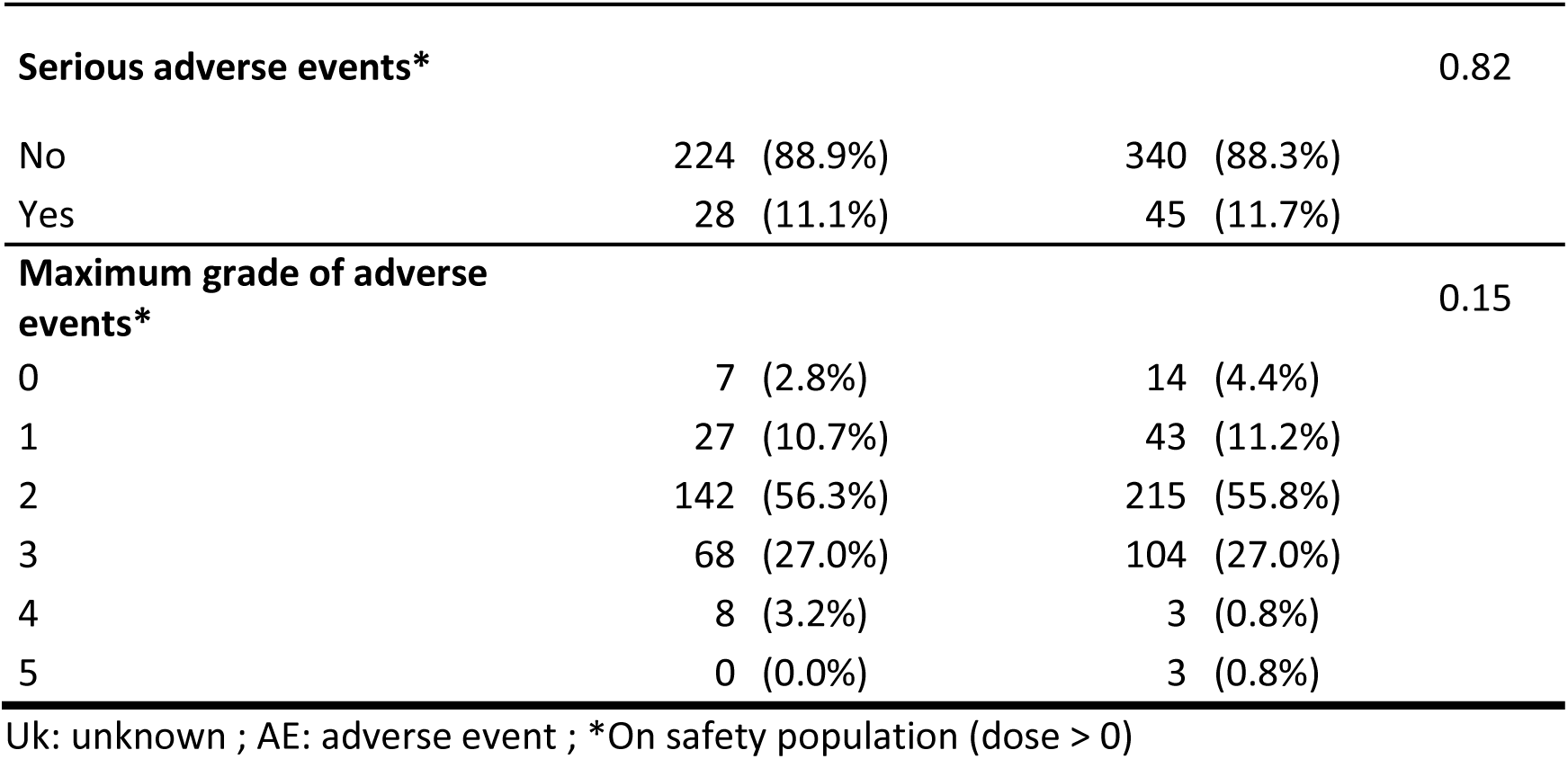
Everolimus dosing and adverse events according to endocrine therapy.

|  | **Tamoxifen**<br>N=252 | **Aromatase inhibitor**<br>N=385 | **P value** |
| --- | --- | --- | --- |
| **Initial everolimus dosing (mg)** |  |  | 0.25 |
| 0 | 2 (0.8%) | 11 (2.9%) |  |
| 2.5 | 0 (0.0%) | 1 (0.3%) |  |
| 5 | 163 (64.9%) | 237 (61.6%) |  |
| 10 | 86 (34.3%) | 136 (35.3%) |  |
| missing | 1 | 0 |  |
| **Early Permanent discontinuation** |  |  | 0.023 |
| No | 88 (34.9%) | 102 (26.5%) |  |
| Yes | 164 (65.1%) | 283 (73.5%) |  |
| **At least one dose reduction** |  |  | 0.90 |
| No | 163 (64.7%) | 251 (65.2%) |  |
| Yes | 89 (35.5%) | 134 (34.8%) |  |
| **At least one AE\*** |  |  | 0.29 |
| No | 7 (2.8%) | 17 (4.4%) |  |
| Yes | 245 (97.2%) | 368 (95.6%) |  |
| **At least one grade 3-5 AE\*** |  |  | 0.67 |
| No | 176 (69.8%) | 275 (71.4%) |  |
| Yes | 76 (30.2%) | 110 (28.6%) |  |
| **When treatment was initiated at 5 mg\*** |  |  | 0.82 |
| No | 122 (75.2%) | 175 (73.8%) |  |
| Yes | 41 (24.8%) | 62 (26.2%) |  |
| **When treatment was initiated at 10 mg\*** |  |  | 0.42 |
| No | 51 (59.3%) | 88 (64.7%) |  |
| Yes | 35 (40.7%) | 48 (35.3%) |  |
| **At least one AE leading to treatment withdrawal\*** |  |  | 0.11 |
| No | 187 (74.2%) | 263 (68.3%) |  |
| Yes | 65 (25.8%) | 122 (31.7%) |  |
| **At least one treatment related AE\*** |  |  | 0.001 |
| No | 17 (6.7%) | 58 (15.1%) |  |
| Yes | 235 (93.3%) | 327 (84.9%) |  |
| **Serious adverse events\*** |  |  | 0.82 |
| No | 224 (88.9%) | 340 (88.3%) |  |
| Yes | 28 (11.1%) | 45 (11.7%) |  |
| **Maximum grade of adverse events\*** |  |  | 0.15 |
| 0 | 7 (2.8%) | 14 (4.4%) |  |
| 1 | 27 (10.7%) | 43 (11.2%) |  |
| 2 | 142 (56.3%) | 215 (55.8%) |  |
| 3 | 68 (27.0%) | 104 (27.0%) |  |
| 4 | 8 (3.2%) | 3 (0.8%) |  |
| 5 | 0 (0.0%) | 3 (0.8%) |  |

Uk: unknown ; AE: adverse event ; \*On safety population (dose > 0)

### Duration of treatment/discontinuation

Despite these data, and most interestingly, early discontinuation of either everolimus or placebo was significantly less frequent in the tamoxifen arm than in the AI arm: 48.0% vs. 56.9% (p=0.028). Accordingly, the median duration of everolimus treatment was significantly longer in the tamoxifen group than in the AI group: 12.8 months (IQR = [2.7-23.6] versus 7.7 months IQR = [1.9-22.6], p=0.007). More specifically, as shown in Figure 3, the 2-year compliance rate with everolimus was approximately 50% in the tamoxifen-treated patients, while it was only 35-40% in the AI-treated patients. Compliance with placebo was similar in both endocrine therapy settings, with a cumulative 2-year compliance rate of approximately 80%. Adherence to therapy is detailed in Table 3. Each row shows adherence as a percentage combining actual therapy duration and dose adjustment. For example, patients in the “<30%” row have taken less than 30% of the theoretical cumulative dose.

**Figure 3.**
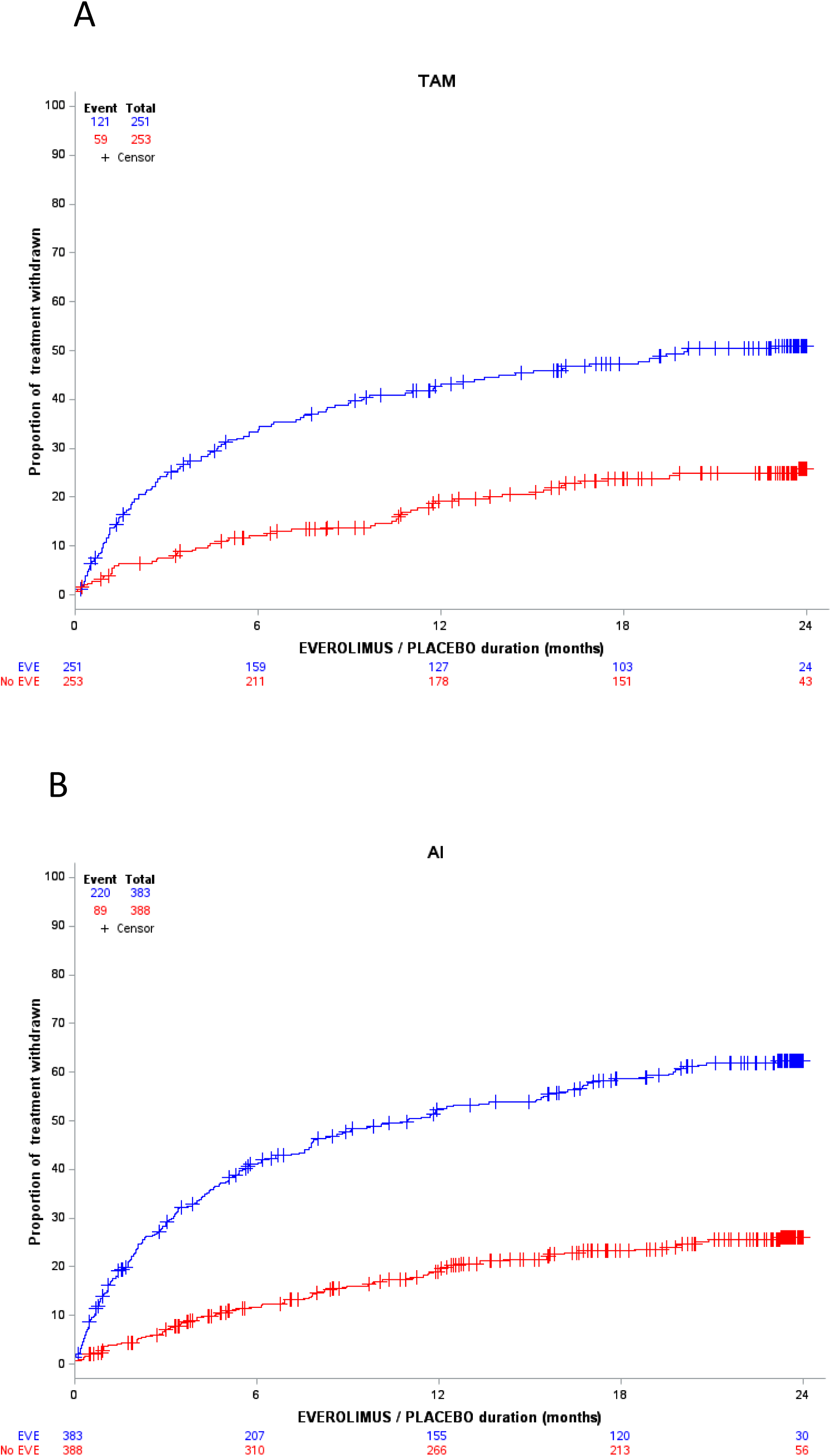
Incidence of Everolimus and Placebo discontinuation according endocrine therapy backbone. Blue curves indicate the everolimus arm; red curves indicate the placebo arm. (A) Patients treated with tamoxifen. (B) Patients treated with an aromatase inhibitor.

**Table 3.** Adherence to therapy in the UNIRAD trial, according to endocrine therapy.

|  | Tamoxifen |  |  |  |  |  | Aromatase inhibitor |  |  |  |  |  |
| --- | --- | --- | --- | --- | --- | --- | --- | --- | --- | --- | --- | --- |
|  | No EVE<br>N=253 (%) |  | EVE<br>N=252 (%) |  | ALL<br>N=505 (%) |  | No EVE<br>N=388 |  | EVE<br>N=385 |  | ALL<br>N=773 |  |
| < 30% | 63 | (24.9%) | 134 | (53.4%) | 197 | (39.1%) | 110 | (28.4%) | 243 | (63.1%) | 353 | (45.7%) |
| [30-50%[ | 41 | (16.2%) | 53 | (21.1%) | 94 | (18.7%) | 71 | (18.3%) | 61 | (15.8%) | 132 | (17.1%) |
| [50-80%[ | 31 | (12.3%) | 29 | (11.6%) | 60 | (11.9%) | 52 | (13.4%) | 48 | (12.5%) | 100 | (12.9%) |
| [80-90%[ | 15 | (5.9%) | 8 | (3.2%) | 23 | (4.6%) | 27 | (7.0%) | 11 | (2.9%) | 38 | (4.9%) |
| [90-95%[ | 21 | (8.3%) | 10 | (4.0%) | 31 | (6.2%) | 31 | (8.0%) | 6 | (1.6%) | 37 | (4.8%) |
| ≥95% | 82 | (32.4%) | 17 | (6.8%) | 99 | (19.6%) | 97 | (25.0%) | 16 | (4.2%) | 113 | (14.6%) |

## Discussion

The addition of everolimus to endocrine therapy is beneficial in patients with advanced ER+ HER2-breast cancer receiving either with tamoxifen^14^, fulvestrant ^17^ or exemestane ^18^. On this basis, and with the goal of improving the outcomes of patients with high-risk early ER+ HER2-breast cancer, we designed the UNIRAD trial to evaluate the addition of 2 years of everolimus to conventional adjuvant endocrine therapy. Unfortunately, the trial did not meet its primary endpoint, likely due to insufficient drug exposure and/or limited activity in this specific clinical setting ^15^. However, pre-specified subgroup analyses suggested a benefit of adjuvant everolimus in patients treated with tamoxifen. This subsequent post-hoc analysis showed a trend in favor of everolimus in the subgroup of premenopausal patients treated with tamoxifen. In general, the most striking difference between patients treated with tamoxifen or an AI was the rate of everolimus discontinuation (48.0% vs 56.9%, respectively) and the duration of exposure to everolimus.

Adverse events associated with everolimus therapy have been widely recognized as an important clinical issue. Dose reductions or discontinuation have been observed in up to 20% of the patients in the metastatic setting, most commonly due to fatigue, rash, diarrhea or stomatitis ^14,17,18^. Of note, the UNIRAD trial was conducted before the generalization of adequate supportive care such as suggested by the SWISH study ^19^. It is also recognized that compliance with endocrine therapy is a major issue in the adjuvant setting, with detrimental consequences for those patients who do not continue treatment ^20^. Notably, these issues of dose reduction/discontinuation and of compliance have also been encountered in recent adjuvant trials evaluating the potential benefit of CDK4/6 inhibitors ^10,21^. For instance, dose reductions were observed in 55% of patients receiving palbociclib in the PALLAS trial ^10^, and in 46% of patients receiving abemaciclib in the Monarch-E trial^8^. However, the dose discontinuation rates were strikingly different (42% in PALLAS and 16.6% in Monarch-E). This may help explain the lack of benefit with palbociclib and the impressive results obtained with abemaciclib, beyond the important differences in the populations of these two studies. Taken together with our present results, these data may partially explain why no benefit was observed with everolimus, at least in patients treated with an aromatase inhibitor.

Conversely, our data suggest that adjuvant everolimus may be beneficial in premenopausal patients treated with tamoxifen. In the SWOG1207 trial, the addition of 1 year of everolimus to endocrine therapy was randomized with no benefit^22^. Strikingly and in line with our results, the unplanned subgroup analyses of this study also identified a potential specific benefit of adjuvant everolimus in premenopausal patients (HR=0.64; 95%CI: 0.44-0.94), and in patients receiving tamoxifen (HR=0.78; 95%CI: 0.53-1.15). Better compliance to treatment may be a first explanation (Figure 3), although no real difference in adverse events was observed between tamoxifen and AI-treated patients. Another important hypothesis is that these patients received suboptimal endocrine therapy, and that everolimus compensated to some extent for this undertreatment. The vast majority of patients receiving tamoxifen in the UNIRAD study were premenopausal (65.7%), and it is noteworthy that ovarian function suppression (OFS) was proposed in only seven of them. At the time the study was designed and conducted, the final results of the SOFT trial, which definitively established OFS as the standard of care for premenopausal patients with high-risk early breast cancer, were not available ^23^, and tamoxifen alone was considered the standard of care for premenopausal women. Since amenorrhea is not a side effect of everolimus ^24^, these results suggest that in this particular setting, everolimus per se may exert a significant clinical activity when added to tamoxifen. Other possible explanations may relate to specific biological interactions between tamoxifen and everolimus. We have found that taking tamoxifen in the evening may be more beneficial than taking it in the morning ^25^. It has also been suggested that patients with menin-low breast cancer may have an improved prognosis when exposed to the tamoxifen-everolimus combination^26^. It may also be hypothesized that everolimus limits activation of the eukaryotic translation initiation factor 4E complex, which has been involved in resistance to tamoxifen^27^.

This report has important limitations. UNIRAD is an underpowered trial, and the present post-hoc exploratory analyses are intended only to generate hypotheses that may help to understand the potential role of everolimus in patients with high-risk early ER+ HER2-breast cancer. We also have not been able so far to explore potential underlying biological differences between premenopausal and menopausal patients.

## Conclusion

Even though current evidence does not support a substantial role for everolimus in the early breast cancer setting ^28,24^, the present data raise interesting hypotheses for the specific subset of premenopausal patients treated with adjuvant tamoxifen. Consistent with current recommendations for adjuvant endocrine therapy, our results also show that tamoxifen alone is suboptimal for high-risk premenopausal patients.

## Data Availability

the CONSORT check list has been thoroughly respected

## Contributions

Conceptualization: MSG, PC, TB

Data curation: Unicancer (JL)

Formal analysis: SC

Funding acquisition: TB, FA, Unicancer (JL)

Investigation: Unicancer

Methodology: SC

Project administration: Unicancer (JL)

Resources: Unicancer (JL) and all authors

Roles/Writing - original draft: PC, MSG, SC, TB

Writing - review & editing: all authors

## Role of the funding source

This trial was supported by a grant from the French Ministry of Health (PHRC 2012) and received funding from La Ligue contre le Cancer, Cancer Research-UK, Myriad Genetics, and Novartis. The finding sources had no role in the whole process of the study or its publication.

**Supplementary Figure. Subgroup analysis of disease-free survival according to age.**

Blue curves indicate the everolimus arm; red curves indicate the placebo arm. (A) DFS in patients <45y old. (B) DFS in patients ≥45y old.

DFS, disease-free survival; HR, hazard ratio; y, years

