## Supplementary figures and images for "Differential benefit of adjuvant everolimus according to endocrine therapy backbone in the randomized UNIRAD trial"

### supplementary figure

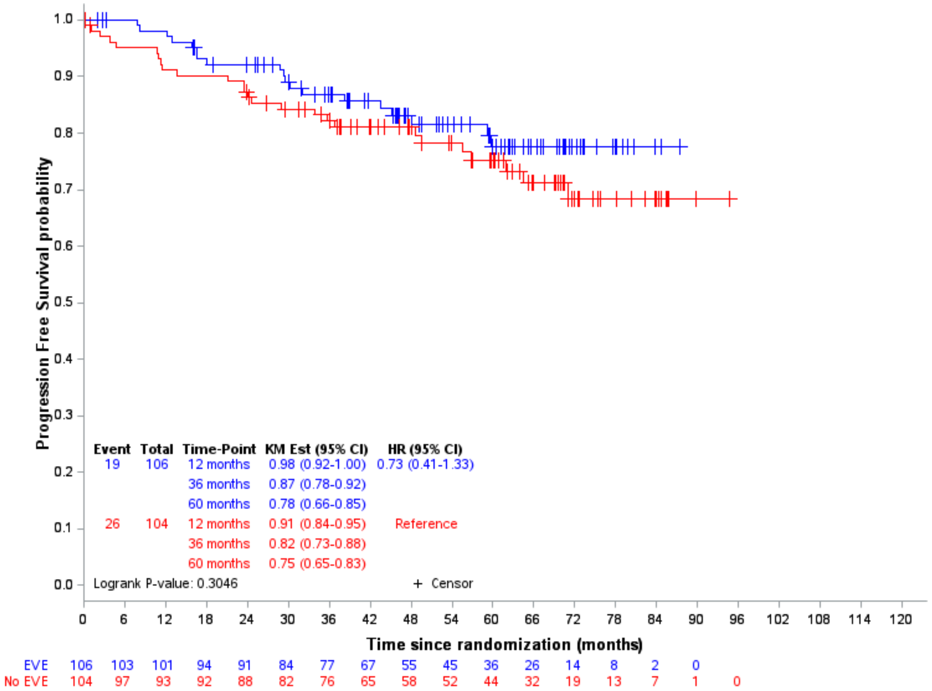

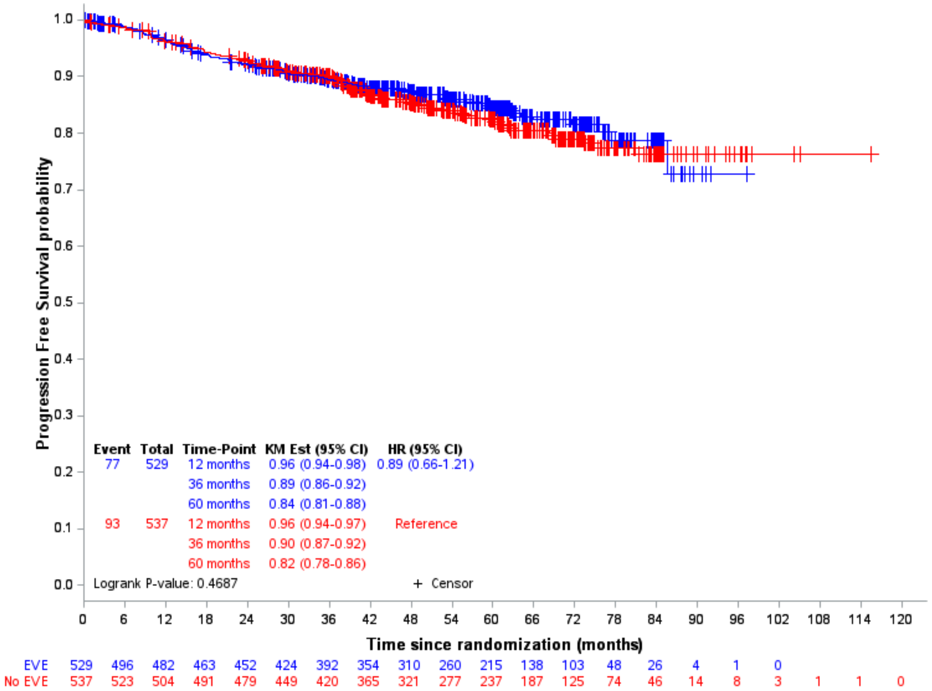
